# Investigating Presence of Ethnoracial Bias in Clinical Data using Machine Learning

**DOI:** 10.1101/2021.09.01.21262949

**Authors:** Bojana Velichkovska, Hristijan Gjoreski, Daniel Denkovski, Marija Kalendar, Leo Anthnoy Celi, Venet Osmani

## Abstract

An important target for machine learning research is obtaining unbiased results, which require addressing bias that might be present in the data as well as the methodology. This is of utmost importance in medical applications of machine learning, where trained models should be unbiased so as to result in systems that are widely applicable, reliable and fair. Since bias can sometimes be introduced through the data itself, in this paper we investigate the presence of ethnoracial bias in patients’ clinical data. We focus primarily on vital signs and demographic information and classify patient ethnoraces in subsets of two from the three ethnoracial groups (African Americans, Caucasians, and Hispanics). Our results show that ethnorace can be identified in two out of three patients, setting the initial base for further investigation of the complex issue of ehtnoracial bias.

## I. Introduction

Machine learning (ML) research is becoming increasingly focused on addressing complex healthcare problems and as a result establishing ML systems for clinical decision support, including diagnostic, prognostic, and risk prediction. In this regard, ML applied to healthcare has already shown significant results [1] [2], which may develop beyond recommendations of clinical actions, towards full-scale assistance such as autonomous triage and patient stratification.

However, the issue of bias in ML research is gaining increasing attention since the results are as reliable as the process is objective. This means that generated results should be unbiased throughout all the stages of the ML process: data collection, data preparation (from data selection to data preprocessing), model configuration, and model training and validation. There are two important aspects to consider: bias can be inherent in the data used in the research or stem from the ML methodology used in the research. Whether the bias is introduced from the data itself or in the development methodology, it presents a significant challenge in terms of trustworthiness of the models and worse can lead to unfair decision making, potentially harming disadvantaged groups, including gender, races and ethnicities.

An important hindrance for increased application of ML models are bias conflicts which must be addressed. An opinion piece [3] states a “silent curriculum” in medical practice teaches students to differentiate between patients based on their race, saying, “*among two patients in pain waiting in an emergency department examination room, the white one is more likely to get medications and the black one is more likely to be discharged with a note documenting narcotic-seeking behavior*”.

Biased medical practice results in ethnoracially unfair medical trials that produce datasets biased towards the majority population, e.g., imbalanced datasets with dominant representation of one ethnorace over the others [4] [5], or datasets obtained entirely from one ethnoracial group. The study in [6] shows that, even though ethnorace influences response to cancer treatments and outcomes, no ethnoracial statuses are recorded in majority of patients, and in cases of recorded ethnorace the highest represented ethnorace in melanoma, breast and lung cancer trials are White people (25.94%), followed by Asians (4.97%), and African Americans (1.08%), resulting in overrepresentation. Additionally, melanoma is one of the deadliest skin cancers known, yet melanoma datasets have shown underrepresentation of different ethnoraces [7].

Working with biased datasets can influence development and produce biased ML applications. There have been many reports of detected racial bias in medical ML applications. The study in [8] shows patients being assigned a risk score depending on their skin color; namely, Black patients which are placed in the same risk category as a subset of White patients, health-wise had considerably worse symptoms. To add to the severity of the problem, the ML algorithm reduced the number of Black patients which should have been assigned additional care by more than half. Another example is an algorithm for diagnosis of diabetic retinopathy showing poor performance in populations living outside of the location where it was developed [9].

Analysis of racial bias in ML applications can also be performed by observing the model’s performance over different ethnoraces [10]. In [11] the authors present their investigation into the performance of three severity scoring systems in four ethnoraces, focusing on hospital mortality; their results show all three models overestimated mortality across all ethnoraces, however, they conclude that severity scores have statistical bias since the overestimated mortalities are most notable with Hispanic and Black patients.

From our investigation it appears there are no existing analysis that focus on detection of racial bias in clinical data itself and hence this is the focus of this paper. Typically, when approaching an ML problem, data preprocessing almost always includes removal of features which could potentially introduce faulty or prejudicial bias in the results, such as gender and ethnorace. However, vital signs, including blood pressure, heart rate and oxygen saturation, are used in clinical data research, and considered unbiased. They are therefore presumed free of information which could be prejudicial in any way. From here, we raise the question, “Is it possible for data, supposedly stripped of biased information, to still incorporate bias in the results?”. Therefore, our aim is finding out if we can detect bias in clinical data, that is, identify ethnicity and race based on vital signs and demographic information only.

The rest of the paper is organized as follows. In section II we describe the dataset, our data preparation process, and our approach to the problem. In section III we present our results, whereas in section IV we discuss them. Section V concludes this paper.

## II. Methodology

### A. Dataset

The dataset used for this paper, eICU Collaborative Research Database (eICU-CRD) [5], contains extensive records of 200,859 patient admissions in the ICU. In this research we focus on the patient’s general information (PGI) and patient’s vital signs measurements (PVS), and are listed along with their units in Table I. During the data preparation process, we consider patients under the age of 89, where all PGI of interest and PVS are present (so called complete case analysis). The PGI we use are patient’s age, height, admission weight, and discharge weight, whereas the PVS we use are statistical features (mean, minimum, maximum, variance, and standard deviation) extrapolated from the patient’s heart rate, oxygen saturation, respiration, and blood pressure (systolic, diastolic, and mean) within the first 24 hours of admission. Patients with undocumented or undefined ethnorace, and patients missing PGI were excluded from this analysis.

**Table I.** Dataset: variables and units

| Variable | Unit |
| --- | --- |
| **Patient’s general information** |  |
| Age | years |
| Height | cm |
| Admission Weight | kg |
| Discharge Weight | kg |
| **Patient’s vital signs** |  |
| Heart Rate | bpm |
| Oxygen Saturation | % |
| Respiratory Rate | insp/min |
| Blood Pressure (systolic, diastolic, mean) | mmHg |

After applying the selection criteria, the dataset contained small pools of patients from certain ethnoracial groups (such as Asian and Native American). Therefore, for our analysis we selected only the three predominant ethnoraces present in the remaining dataset, namely Caucasian, African American and Hispanic. The distribution of patients per ethnoracial group is given in *Fig. 1*. As the chart shows, the resulting dataset is highly imbalanced, in favor of Caucasians. The African American and Hispanic ethnoraces have a significantly lower number of patients, with the Hispanic group being the smallest.

**Fig. 1.**
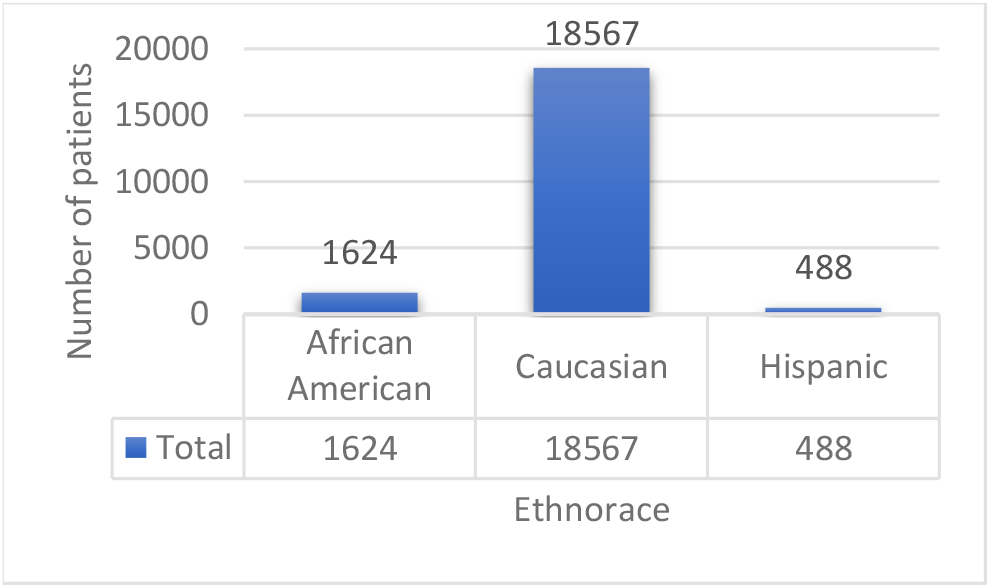
Patient number per ethnorace

### B. Model development and validation

We classify patients’ ethnoraces in subsets of two from the three ethnoraces we have, i.e., we have three distinct comparative tests, Caucasian vs African American, African American vs Hispanic, and Caucasian vs Hispanic patients. For each comparative test, we evaluate the performance of two ML algorithms, namely logistic regression (LR) and XGBoost. LR predicts the probability of dichotomous target variables. XGBoost is an efficient implementation of the gradient boosted trees, which makes predictions by ensembling new models to correct the errors made by existing models, until no further improvements can be made. The evaluation of the models is performed internally using stratified five-fold cross validation. In five-fold cross validation, four folds are used for the model training, whereas the remaining fold is used for testing the model’s performance. We use stratified cross validation in order to maintain the original classes’ distribution.

The assessment of our models is performed by computing the area under the receiver operator characteristic curve (AUC-ROC), the area under the precision-recall curve (AU-PRC), and additional metrics, including positive predictive value (PPV), negative predictive value (NPV), F1 score, and recall for each model. The AUC-ROC curve shows the trade-off between true positive rate (TPR) and false positive rate (FPR). TPR (also known as recall and sensitivity) is the proportion of samples correctly predicted as positives out of all positive observations. FPR is the proportion of samples incorrectly predicted as positives out of all negative observations. Classifiers with curves closer to the top-left corner have better performance compared to classifiers with a curve closer to the 45-degree diagonal. The AU-PRC shows the trade-off between precision or PPV and TPR. PPV represents the proportion of samples predicted correctly as positives out of all samples predicted as positives. NPV represents the proportion of samples correctly predicted as negatives and all samples predicted as negatives. F1 score is the harmonic mean of precision and recall.

## III. Experimental Results

Table II shows the difference between each pair of ethnoraces (classes) in our dataset. The ratios provided are important indicators when analyzing the performance of our models, since the AU-PRCs obtained are influenced by data ratio. We can see that the highest data imbalance occurs between Caucasians vs Hispanics, whereas the lowest data imbalance can be seen in African Americans vs Hispanics.

**Table II.** Patient number ratio between majority and minority classes in each comparative test

| Dataset: Class Distribution |  |  |
| --- | --- | --- |
| Majority class | Minority class | Patient Number Ratio |
| Caucasian | African American | 11.42 |
| African American | Hispanic | 3.32 |
| Caucasian | Hispanic | 37.90 |

Our results consist of two sets: first, using imbalanced data in the training process, and second, using balanced data in the training process. In both cases we maintained the original class ration for the test data.

### A. Imbalanced train data

Initially, we used an imbalanced dataset, both for training and testing the models. All three comparative tests showed the model was biased in favor of the majority class. Since the ratio between African Americans and Hispanics is the lowest imbalance ratio in our dataset, we decided to show the performance of the ML models trained on the original distribution of data from these two classes.

Table III shows confusion matrices taken from a random fold for LR [12] and XGBoost [13], and for both classifiers most of the patients are classified as part of the majority class. Therefore, these results clearly illustrate that the imbalance in the data made the models biased towards the majority class. Caucasian patients are 11.42 times more than African American patients and 37.9 times more than Hispanics, which further accentuates the bias towards Caucasians, when using the imbalanced dataset for training.

**Table III.** African American vs Hispanic - confusion matrices. Taken from a randomly selected fold.

| African American vs Hispanic |  |  |  |  |
| --- | --- | --- | --- | --- |
| Models | Logistic Regression |  | XGBoost |  |
|  | Predicted |  |  |  |
| Actual | 316 | 9 | 304 | 21 |
|  | 96 | 2 | 87 | 11 |

### B. Balanced train data

Since the imbalanced dataset proved to be biased towards the majority class and resulted in models placing most of the samples in the dominant class, understandably there was low model performance. In order to give the models a learning chance, we decided to correct the imbalance present in the dataset, during the training process. To achieve this, we randomly under sampled the majority class (using RandomUndersample [14]) in the training data, as to balance the train dataset. We train the classifiers with the balanced data, while the test data for each fold keeps the original distribution in order to evaluate the performance of each model on real data.

Using the balanced train data, we illustrate results from three comparative binary classifications between each two ethnoracial groups – firstly, we have Caucasians vs African Americans, next we have African Americans vs Hispanics, and lastly, we have Caucasians vs Hispanics.

The results of each comparative test for each classifier are summarized in Table IV, Table V, and Table VI. 95% confidence intervals are provided in the brackets. Additionally, the AUC-ROC and AU-PRC are illustrated in Fig. 2, where the 95% confidence intervals for the classifiers are shown in their corresponding coloring with lower opacity.

**Table IV.** Caucasian vs African American - results. Confidence intervals provided in brackets.

| Metric | LR | XGBoost |
| --- | --- | --- |
| AUC | 0.583 [0.552 – 0.614] | 0.726 [0.703 – 0.749] |
| PPV | 0.121 [0.114 – 0.128] | 0.138 [0.134 – 0.142] |
| Recall | 0.558 [0.501 – 0.615] | 0.640 [0.599 – 0.681] |
| NPV | 0.944 [0.938 – 0.950] | 0.954 [0.950 – 0.958] |
| F1 | 0.199 [0.186 – 0.212] | 0.227 [0.220 – 0.234] |

**Table V.**
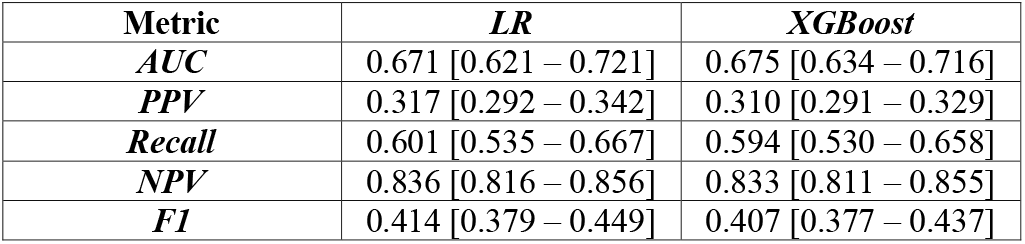
African American vs Hispanic - results. Confidence intervals provided in brackets.

**Table VI.** Caucasian vs Hispanic - results. Confidence intervals provided in brackets.

| Metric | LR | XGBoost |
| --- | --- | --- |
| AUC | 0.737 [0.693 – 0.781] | 0.648 [0.636 – 0.660] |
| PPV | 0.045 [0.040 – 0.050] | 0.044 [0.043 – 0.045] |
| Recall | 0.602 [0.524 – 0.680] | 0.627 [0.616 – 0.638] |
| NPV | 0.985 [0.982 – 0.988] | 0.985 [0.984 – 0.986] |
| F1 | 0.083 [0.073 – 0.093] | 0.081 [0.080 – 0.082] |

**Fig. 2.**
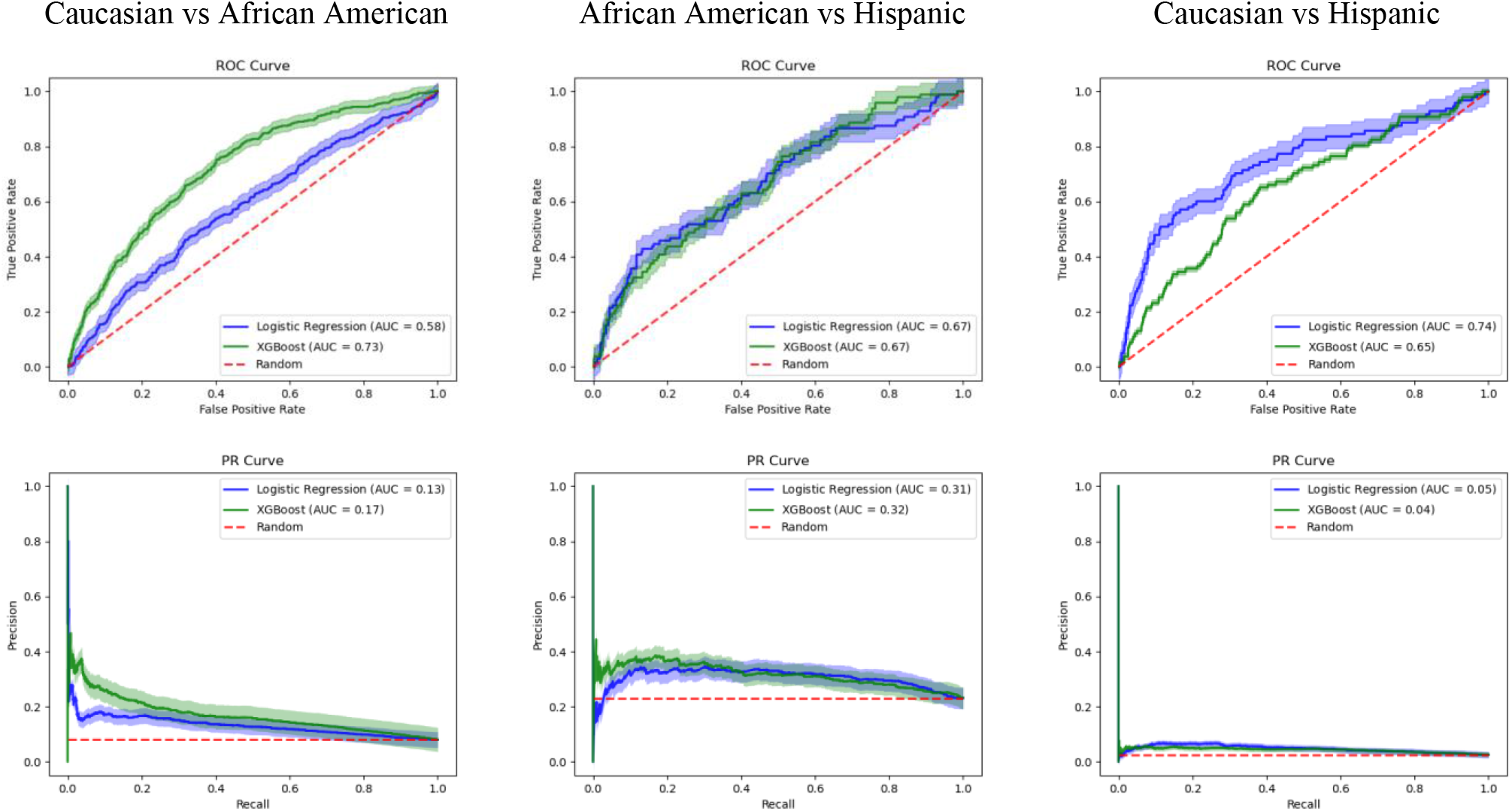
Performance on each model across three comparative tests. The top row represents AUC-ROC, the bottom row represents AU-PRC. Confidence intervals are shown with lower opacity.

## IV. Discussion

From the results, it can be observed that XGBoost performed best in classifying Caucasians vs African Americans, whereas the other two comparative tests give weaker results. For both African Americans vs Hispanics and Caucasians vs Hispanics, XGBoost shows significant similarity in the AUC-ROC curve. On the other hand, LR has the worst performance for Caucasians vs African Americans, and the best performance for Caucasians vs Hispanics. This outcome is understandable, because LR is known to operate well even with small sample sizes, which is the case in the last comparison. However, from the confidence intervals for both classifiers along all comparisons we can see that XGBoost has a narrower range around the estimate, which means that the estimate provided by XGBoost is more stable compared to the estimate given by LR.

Observing the additional metrics obtained in the experiments we can see that throughout all of them the confidence intervals are narrower for XGBoost.

The PPVs for all the experiments are low. However, the values for the recall (which show us the number of correctly returned patients divided by the number of patients which should have been returned) show that on average two thirds of patients are correctly classified, as is further illustrated with the randomly selected confusion matrices provided in Table VII.

**Table VII.**
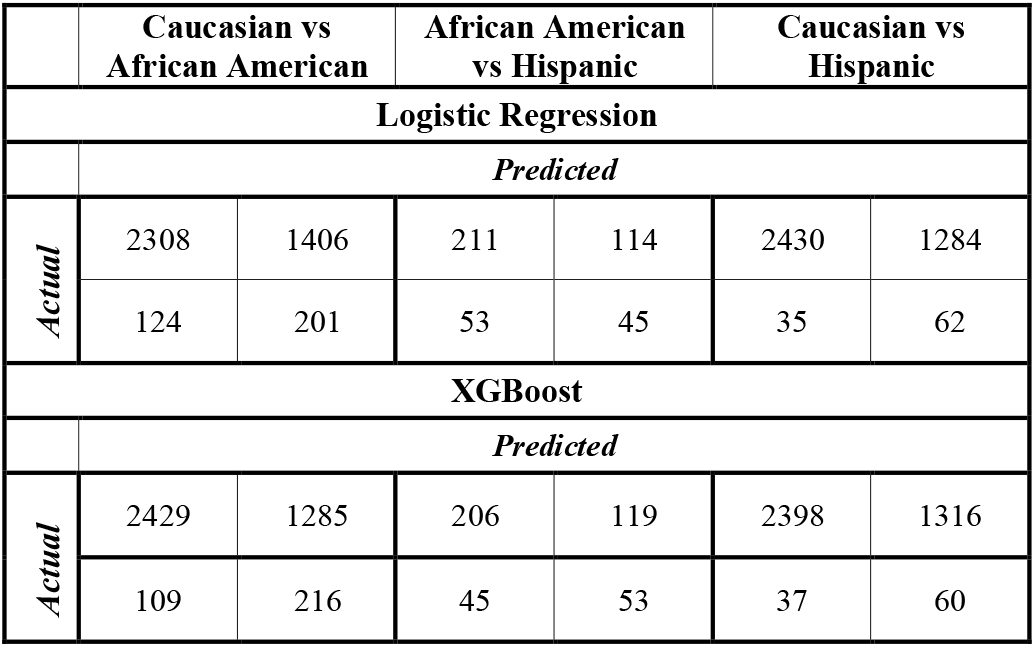
Confusion matrices for all experiments. Taken from a randomly selected fold.

These results show that patient’s general information and vital signs could include ethnoracial bias. Perhaps this bias arises from the bias present in medical practice, e.g., Black or Hispanic patients admitted to the ICU might be in a worse condition than White patients. Another potential reason can be similarities in general information and biological markers (e.g., height, weight, heart rate) of patients that represent an ethnoracial group.

However, these results are not conclusive. Ethnoraces can be difficult to identify due to interracial marriages, and we cannot claim with certainty that Caucasians “misclassified” as African Americans, are not biracial or even multiracial. Furthermore, the eICU dataset consists of patients with various diagnosis, e.g., rhythm disturbances, pneumonia, aneurysms, and so on, and every diagnosis influences different vital signs in different ways. Additionally, the results are obtained on a small number of African American and Hispanic patients, which might not give an accurate representation on these ethnoracial groups.

## V. Conclusion

With the increased number of ML applications in medicine it is important to ensure the developed models are unbiased and perform correctly in spite of a patient’s ethnorace. Since bias can be introduced through data, we investigated the presence of ethnoracial bias in clinical data; more specifically, we analysed general information and vital signs of patients from three ethnoraces to determine whether ML models can detect biological markers representative of an ethnorace. We compared the performance of two ML algorithms in comparing two by two ethnoraces in balanced train data. Our results show that two out of three patients in all experiments are placed in the correct ethnorace; however, the sample size of the observed ethnoraces as well as the fluid concept of ethnorace indicate the need for further investigation.

## Data Availability

The datasets analyzed in the current study are publicly available in the eICU-CRD repository.

https://eicu-crd.mit.edu/

## Declarations

### Ethical approval and consent to participate

The analysis using the eICU-CRD is exempt from institutional review board approval due to the retrospective design, lack of direct patient intervention, and the security schema, for which the re-identification risk was certified as meeting safe harbor standards by an independent privacy expert (Privacert, Cambridge, MA) (Health Insurance Portability and Accountability Act Certification no. 1031219-2). All experiments were performed in accordance with relevant guidelines and regulations.

### Consent for publication

Not applicable.

### Availability of data and materials

The datasets analyzed in the current study are publicly available in the eICU-CRD repository (https://eicu-crd.mit.edu/).

## Acknowledgements

Not applicable.

